# Real-world evidence of mortality and survival rates in 256 individuals with APDS

**DOI:** 10.1101/2022.12.05.22283110

**Authors:** Jennifer Hanson, Penelope E. Bonnen

## Abstract

Activated Phosphoinositide 3-kinase Delta Syndrome (APDS) is a rare genetic disorder that presents clinically as a primary immunodeficiency. Clinical presentation of APDS includes severe, recurrent infections, lymphoproliferation, lymphoma and other cancers, autoimmunity and enteropathy. Autosomal dominant variants in two independent genes have been demonstrated to cause APDS. Pathogenic variants in *PIK3CD* and *PIK3R1*, both of which encode components of the PI3-kinase, have been identified in subjects with APDS. APDS1 is caused by gain of function (GOF) variants in the *PIK3CD* gene while loss of function (LOF) variants in *PIK3R1* have been reported to cause APDS2. We conducted a review of the medical literature and identified 256 individuals who had a molecular diagnosis for APDS as well as age at last report; 193 individuals with APDS1 and 63 with APDS2. A Kaplan-Meier survival analysis for APDS showed the conditional survival rate at the age of 20 was 87%, age 30 was 74%, age 40 and 50 were 68%. Review of causes of death showed that the most common cause of death was lymphoma, followed by complications from HSCT. The mortality data suggests that the standard of care treatment for APDS, immunoglobulin replacement therapy, appears to prevent most deaths due to severe infection, however, new treatments are needed to mitigate the risk of death from lymphoma and other cancers. This analysis based on real world evidence gathered from the medical literature is the largest study of survival for APDS to date.

## Introduction

Activated Phosphoinositide 3-kinase Delta Syndrome (APDS) is a rare genetic disorder that presents clinically as a primary immunodeficiency. Autosomal dominant variants in two independent genes have been demonstrated to cause APDS. Pathogenic variants in *PIK3CD* and *PIK3R1*, both of which encode components of the PI3-kinase, have been identified in subjects with APDS. APDS1 is caused by gain of function (GOF) variants in the *PIK3CD* gene, while loss of function (LOF) variants in *PIK3R1* have been reported to cause APDS2 ^1-4^. The pathogenic variants *PIK3CD* and *PIK3R1* result in the over activation of PI3-kinase. PI3-kinase plays a role in regulation of T and B cells, therefore, both APDS1 and APDS2 manifest as combined immunodeficiency. Clinical presentation of APDS typically begins in the first year of life as severe, recurrent infections. This progresses to include lymphoproliferation and sometimes malignant lymphoma; many patients also experience autoimmunity and enteropathy ^5^.

Treatment for APDS often includes immunoglobulin replacement therapy to combat recurrent infection, as well as immunosuppressive agents such as rituximab, sirolimus and tacrolimus to mitigate autoimmunity and lymphoproliferation ^5^. Hematopoietic stem cell transplantation (HSCT) has also been undertaken in a minority of patients and has been shown to ameliorate symptoms, however, HSCT itself can also cause adverse complications and death ^5-8^.

Despite available treatments, survival for individuals with APDS appears to be shortened from the average lifespan ^6; 7^. Okano *et al* reported on APDS1 survival based on 23 Japanese and Taiwanese patients from 21 families, 9 of which had HSCT ^6^. Thirty-year survival was 83% with just 2 deaths, both due to HSCT. Elkaim *et al* reported that the thirty-year survival rate of APDS2 was 83%, based on an international cohort of 36 patients ^7^.

We conducted a review of the medical literature for every published case of APDS with documented age at last report and a molecular diagnosis in *PIK3CD* or *PIK3R1*. We identified 256 individuals who had a molecular diagnosis for APDS as well as age at last report; 193 individuals with APDS1 and 63 with APDS2. A comprehensive survival analysis and study of causes of death for APDS was conducted. The results of this study show decreased survival compared to previous reports, which may be attributed to the sample size of this study being more than 7x higher than prior studies.

## Methods

### Literature review and data extraction

Literature was reviewed for all reports of age of individuals with APDS. All individuals included in this analysis were published in English language peer-reviewed journals indexed in PubMed. Search terms utilized were PIK3CD, PIK3R1, PASLI, APDS. In order to be included in this study an individual had to have been reported to have a molecular diagnosis of pathogenic variant in either PIK3CD or PIK3R1. Age at last report was an additional requirement to be included in this study. In addition, cause of death and wether the individual received HSCT were also noted when available. In total, 116 papers were reviewed. Last access date was 08-01-2022. Through this review of the medical literature, 193 individuals diagnosed with APDS1 through molecular genetic testing and pathogenic variant present in *PIK3CD* were extracted from the literature for whom age was reported. In addition, 63 individuals diagnosed with APDS2 through molecular genetic testing and pathogenic variant present in *PIK3R1* were extracted from the literature for whom age was reported.

### Kaplan-Meier survival analysis

Kaplan-Meier survival analysis was conducted using the R survival package to estimate the probability of survival over time plus the 95% confidence interval ^9-11^. All-cause mortality was considered the endpoint. Any individual who was alive at last report was censored. A censored observation is one where the subject drops out of the study but survives at a given time point. Kaplan-Meier survival analysis was conducted using 1-year age bins from birth to the oldest patient age. The Mantel-Haenszel test was utilized to test if the conditional probability estimates from APDS1 and 2 were different from each other ^12^.

## Results

We reviewed the literature for all reports of age of individuals with APDS. Through this review of the medical literature, 193 individuals diagnosed with APDS1 through molecular genetic testing and pathogenic variant present in *PIK3CD* were extracted from the literature for whom age was reported ^1; 2; 6; 8; 13-54^. In addition, 63 individuals diagnosed with APDS2 through molecular genetic testing and pathogenic variant present in *PIK3R1* were extracted from the literature for whom age was reported ^3; 4; 7; 8; 26; 44; 55-66^.

The median age of individuals reported with APDS1 was 13 years with an average of 17 years (range, 1 - 64 years). The median age of individuals reported with APDS2 was 14 years with an average of 16 years (range, 1 - 56 years). The age distribution was significantly different between the two groups due to there being disproportionately more individuals with APDS1 in the 5 – 15 year age range (p = 3.6E-10) (Figure 1). Gender was not available for all individuals in the study. For APDS1, 103/182 (57%) individuals were reported as being male and 79/182 (43%) individuals were noted to be female. For APDS2, 20/39 (51%) individuals were reported as being male with 19/39 (49%) individuals were noted to be female.

**Figure 1.**
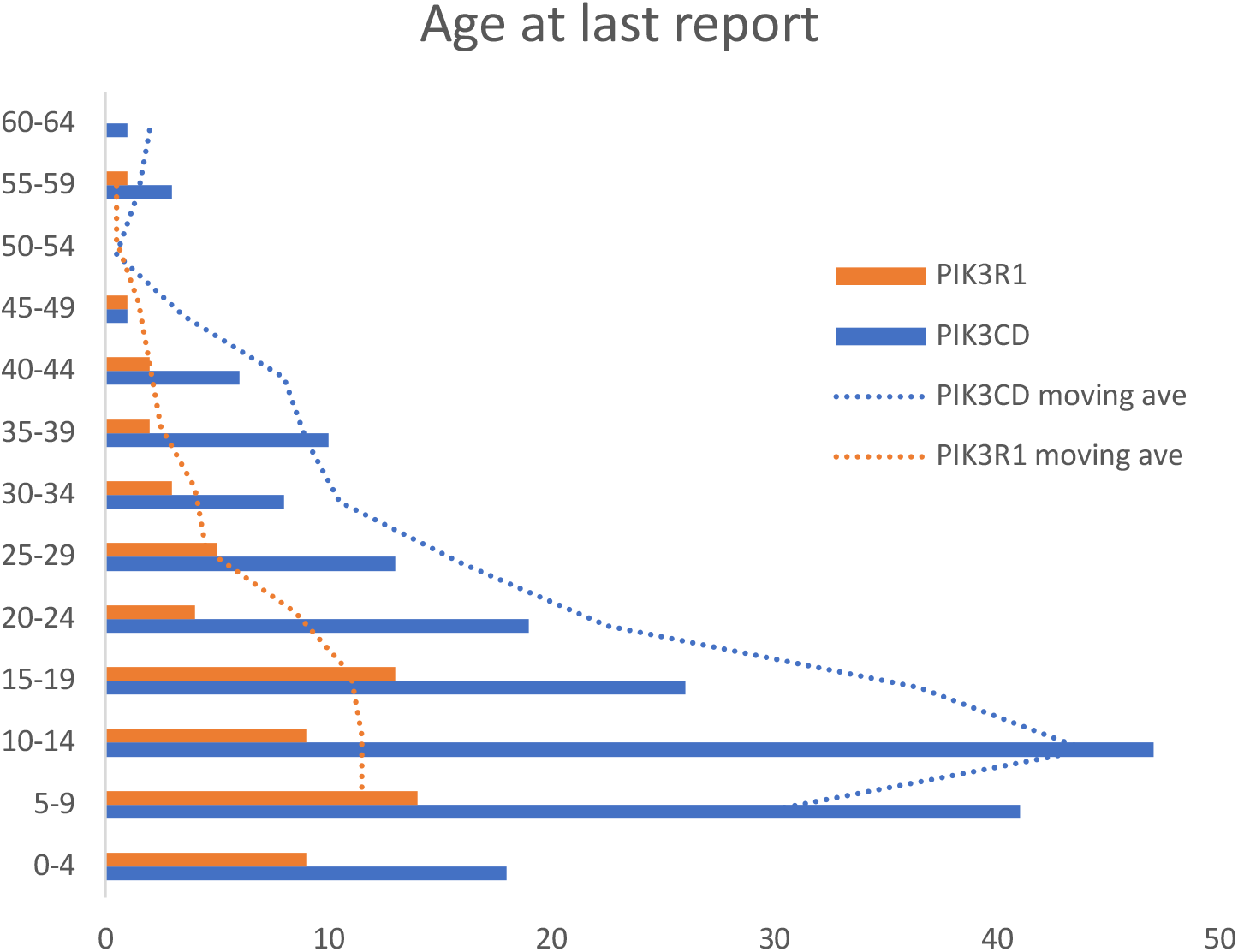
Age at last report in a cohort of 256 individuals with APDS. The age at last report was plotted in 5-year age bins for individuals with a molecular diagnosis of pathogenic PIK3CD variant resulting in APDS1 (orange), and for those with a molecular diagnosis of pathogenic PIK3R1 variant (blue). The moving average of the age at last report was plotted for individuals with APDS1/PIK3CD in blue and for individuals with APDS2/PIK3R1 in orange.

Twenty-four individuals with APDS1 in this study were reported as being deceased. Age of death ranged from 1 - 64 years. The most common cause of death was tied between lymphoma (N = 5) and HSCT (N = 5) (Figure 2). The range of ages for death by lymphoma was 1 – 27 years, while the range of ages for death by HSCT was 5 – 18 years. The next most common cause of death was sepsis with no further information specified (N = 3, age range 11 – 31 years). Additional causes of death included: Varicella zoster pneumonitis (N = 1, age 12); Acute myeloid leukemia (N = 1, age 22); lymphoproliferative disease (N = 1, age 11); gastric cancer (N = 1, age 64); IgA nephropathy (N = 1, age 57); respiratory failure (N = 1, age 39) (Figure 2).

**Figure 2.**
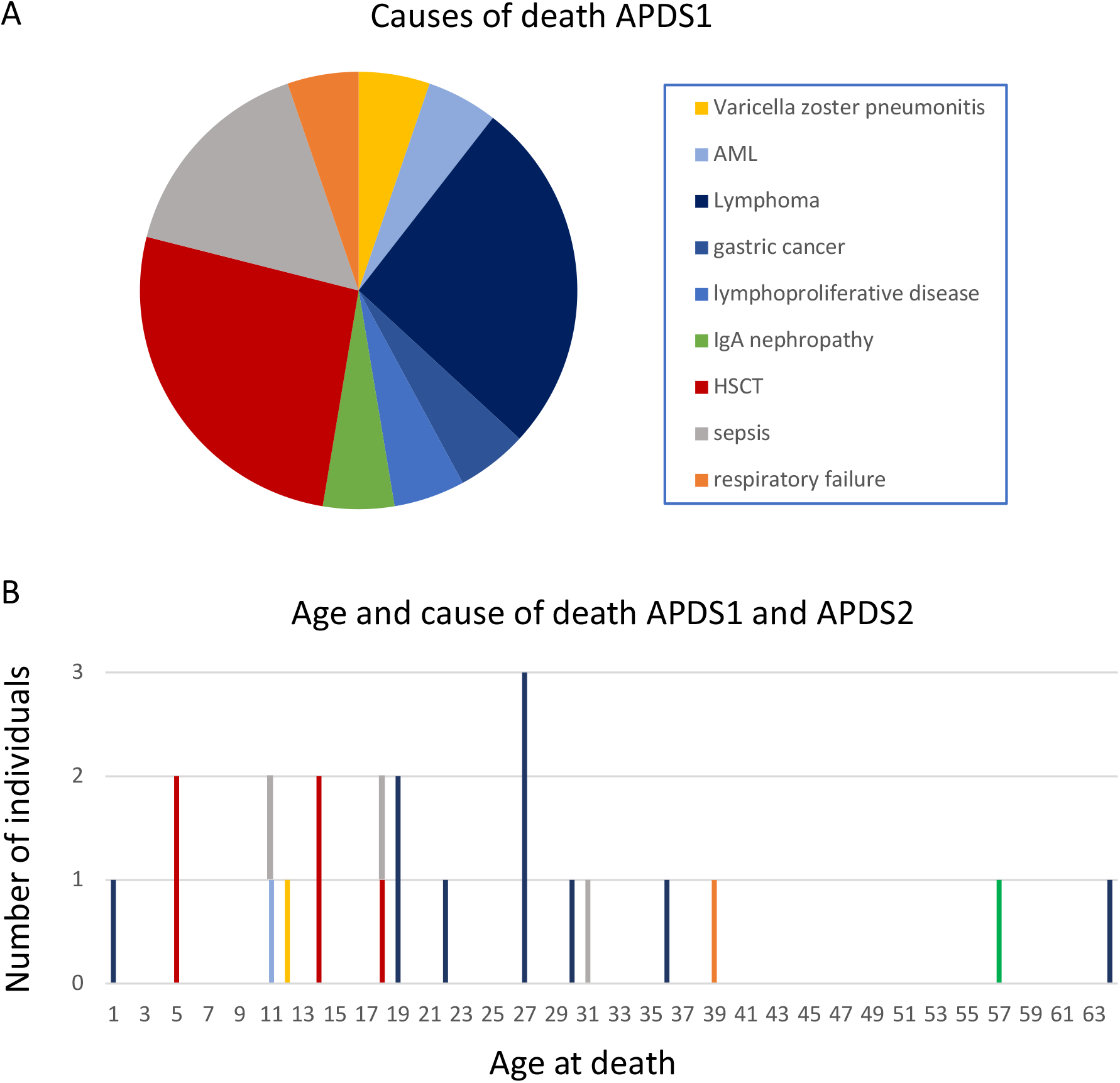
Age and cause of death for APDS. A. Causes of death in individuals with APDS1 are shown and the size of the pie slice reflects the number of individuals with that cause of death. All individuals with APDS2 had the same cause of death, lymphoma, so it was not plotted. B. Cause of death in individuals with APDS1 and APDS2 was plotted with age of death on the x-axis and number of individuals plotted on the y-axis. The color scheme in A. and B. are the same, with each color indicating a different cause of death. The key for color to cause of death is shown in the figure.

Five out of the 63 individuals with APDS2 in this study were reported as having died. Age of death ranged from 12 - 36 years. The cause of death was reported for 4/5 individuals who died with APDS2 and all four of their deaths were attributed to lymphoma.

Kaplan-Meier survival analysis was conducted that allowed for inclusion of individuals who were alive at last report as well as those who died during the timespan of the study. Kaplan-Meier survival analysis was conducted for all-cause mortality in 1-year age bins on subjects diagnosed with APDS1 and separately for subjects diagnosed with APDS2 (Figure 3). The survival probability estimate for APDS1 at the age of 20 was 87%, age 30 was 75%, age 40 and 50 was 69%. The survival probability estimate for APDS2 at the age of 20 was 98%, age 30 was 72%, age 40 and 50 was 60%. The Mantel-Haenszel test showed that the survival probability estimates for APDS1 and APDS2 were not different from each other (p-value: 0.49). Consequently, Kaplan-Meier survival analysis was also conducted on the combined APDS1-2 cohort using age at last report for 256 individuals, since the combined cohort was largest and had the most power (Figure 3). Because most of the individuals in the combined cohort had APDS1 the survival curve for APDS1-2 most closely reflected the survival curve for APDS1. For APDS1-2 the conditional survival rate at the age of 20 was 87%, age 30 was 74%, age 40 and 50 was 68%.

**Figure 3.**
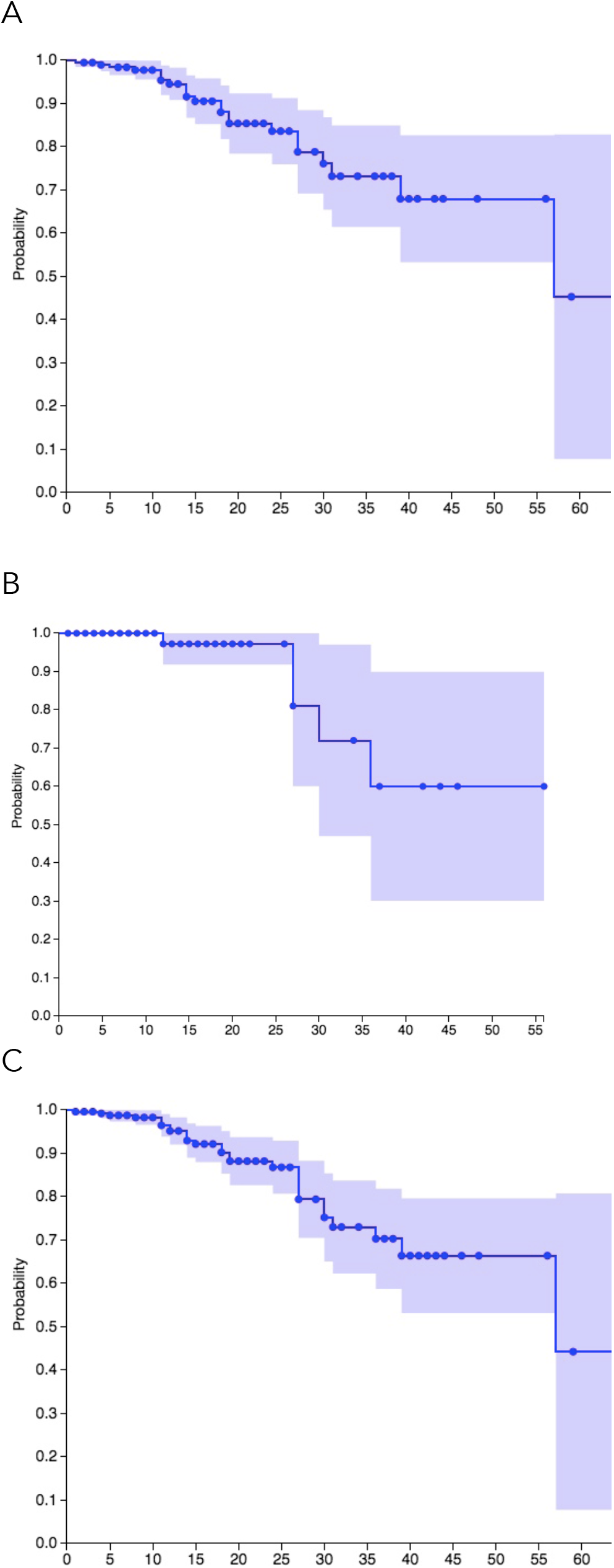
Kaplan-Meier survival analysis of all-cause mortality in 256 individuals with APDS. A. The probability of survival over time plus the 95% confidence interval was plotted for individuals with APDS1, N = 193, and separately for B. APDS2, N = 63. The survival rate conditional on having either APDS1 or APDS2, N = 256, was estimated by Kaplan-Meier analysis and plotted in panel C.

## Discussion

Kaplan-Meier survival analysis and studies of mortality was conducted for 256 individuals who had genetically defined APDS1 or 2. Kaplan-Meier survival analysis on the combined APDS1-2 cohort using age at last report showed that the APDS1-2 the conditional survival rate at the age of 20 was 87%, age 30 was 74%, age 40 and 50 was 68%.

In this cohort of 256 individuals with APDS1 or APDS2, 29 deaths were noted. Of these, the cause of death was reported for 23 individuals. The most common cause of death was lymphoma with age of death from lymphoma ranging from 1 – 27 years. The second most common cause of death was complications resulting from HSCT. Of the individuals who reached older ages, the causes of death were gastric cancer (age 64), IgA nephropathy (age 57), and respiratory failure (age 39). Most deaths occurred before the age of 30, however, it is important to note that the age distribution of this cohort is highly skewed toward younger ages with 85% of the combined APDS cohort in this study being less than 30 years old.

Given that only 15% of the cohort was older than 30, this study has less power beyond the age of 30. This is reflected in the larger confidence intervals in the Kaplan Meier curves post-30, as well as the dearth of deaths after age 30 in the chart in Figure 2B. The chart could be misconstrued to indicate that there are fewer deaths after the age of 30, however, the data from this cohort simply does not provide the opportunity to make observations post-age 30 with as much sensitivity as below age 30. Having noted this limitation, this cohort contained 38 individuals 30 years or older which is larger than the total study size of the two previous survival analyses published for APDS.

There are two previously published survival analyses for APDS, one for APDS1 and one for APDS2 ^6; 7^. With much smaller sample sizes than this study, just N = 23 for APDS1 ^6^ and N = 36 for APDS2 ^7^, both reported thirty-year survival was 83%. In this study with a much larger cohort, N = 256, the thirty-year survival was 74%. The lower survival rates in this study is likely due to the larger sample size of this cohort resulting in greater sensitivity to observe events and consequently in a more robust measure of survival rates.

Common variable immune deficiency (CVID) is a heterogenous group of primary immune deficiencies clinically-defined as having reduced IgG, IgA, and/or IgM. There are multiple genetic causes of CVID with *PIK3CD* and *PIK3R1* being among them ^67^. Kaplan-Meier survival analysis of a large cohort of 411 subjects with genetically undefined CVID who were followed for four decades was reported ^68^. The 30-year survival rate for this CVID cohort was 68% for females and 70% for males. This more closely resembles the 74% thirty-year survival rate that was found in this study of APDS.

The most common causes of death in the CVID cohort were respiratory failure from chronic lung disease (37%), cancer (primarily lymphoma) (29%), and severe infection (10%) ^68^. In contrast, lymphoma was the most common cause of death in this APDS cohort and only one person with APDS was reported as having died due to respiratory failure from chronic lung disease. This could indicate that respiratory failure is a less frequent feature of APDS than heterogenous CVID; however, it could also be that respiratory failure from chronic lung disease occurs at older ages, and as we noted this APDS cohort has 38/256 individuals who were older than 30. Longer term follow-up on APDS is warranted to better define survival and mortality at older ages.

The results of this study on survival and mortality in APDS suggest that the standard of care treatment for APDS, replacement immunoglobulin therapy, appears to prevent most deaths due to severe infection, however, new treatments are needed to mitigate the risk of death from lymphoma and other cancers.

## Data Availability

All data produced in the present study are available upon reasonable request to the authors

